# Leveraging probabilistic forecasts for dengue preparedness and control: the 2024 Dengue Forecasting Sprint in Brazil

**DOI:** 10.1101/2025.05.12.25327419

**Authors:** Eduardo Correa Araujo, Luiz Max Carvalho, Fabiana Ganem, Luã Bida Vacaro, Leonardo S. Bastos, Laís Picinini Freitas, Marcio Bastos, Ramila Alencar, Lucas Bianchi, Raúl Capellán, Xiang Chen, Oswaldo Cruz, Americo Cunha, Haridas K. Das, Chloe Fletcher, Raquel Martins Lana, Rachel Lowe, Daniela Lührsen, Giovenale Moirano, Paula Moraga, Lucas M. Stolerman, Fernanda Valente, Cláudia Torres Codeço, Flávio C. Coelho

## Abstract

Forecast models are a key decision-support tool for public health authorities in managing epidemics, feeding into early warning systems, scenario evaluations, and empirical basis for resource allocation. In Brazil, improving dengue forecasting became a priority in response to the unprecedented increase in cases, which surpassed the total of the previous decade and expanded to new regions. The Infodengue-Mosqlimate consortium launched the Brazilian Dengue 2024 Challenge (IMDC24), or Dengue Forecast Sprint, bringing together six international teams provided with cases and climate covariates data to generate actionable forecasts for 2024 and 2025 seasons in five diverse Brazilian states, leveraging advanced machine learning and classical statistical models. This paper outlines the structure and findings of the IMDC24. Model performance varied between years and locations, and no single model consistently excelled, especially during 2024’s atypical, climate-change-driven conditions. This performance variability highlighted the need for ensemble approaches. The ensemble models developed are presented as the main results of this collaborative development. As intended, the ensemble models have been adopted by Brazilian public health authorities to help with planning and response to the forecasted 2025 dengue epidemics across the country.

**Significance Statement:** The Dengue Challenge 2024 (IMDC24) was organized by the Mosqlimate-Infodengue consortium, which aims to provide forecasting models as decision support tools for early warning systems, scenario assessments, and empirical basis for resource allocation for mosquito-borne diseases. During IMDC24, six international teams, provided with dengue case, sociodemographic, and climate data, developed scenario forecasting models for the 2024 and 2025 dengue seasons in Brazil. In this study, we evaluated the performance of each model and built an ensemble model, considering the variation in performance of each model, especially during the atypical climate conditions of 2024. Among the main applications of this work, we highlight the incorporation of the results into the Brazilian Ministry of Health’s nationwide dengue epidemic response agenda.

## Introduction

Forecast models are a key asset in the response to epidemics by public health authorities, informing their control strategies [Bicher et al., 2022, Viboud et al., 2018, Mathis et al., 2024]. Output from these models can provide early warning of emerging outbreaks, help with the evaluation of scenarios, and enhance the precision of resource allocation. Developing an appropriate forecasting model depends on specific research questions and public health objectives. Each model has unique strengths and weaknesses, and a single model is unlikely to encapsulate all relevant aspects of disease dynamics. For instance, while deep learning models are adept at capturing temporal structures and nonlinearities, classical statistical models like SARIMA can perform comparably well, especially in systems with pronounced seasonal patterns such as those observed in dengue. This underscores the need for rigorous benchmarking and model comparison using appropriate evaluation metrics across diverse modeling domains [Mills et al., 2024].

Numerous epidemic forecast initiatives have been proposed [Reich et al., 2019a, Johansson et al., 2019, Reich et al., 2019b, Cramer et al., 2022, Bracher et al., 2021, Funk et al., 2020, Mathis et al., 2024], including probabilistic superensemble models that integrate seasonal climate forecasts and lagged disease cases [Colón-González et al., 2021] and ensemble machine learning methods that include advanced deep neural networks [Sebastianelli et al., 2024, Baquero et al., 2018]. Moreover, it is also crucial to combine predictions from various models in *ensembles*, that capture the main features of each model while guarding against overconfidence or catastrophic failure [Gneiting and Raftery, 2005, Johansson et al., 2019, Reich et al., 2019b, Viboud et al., 2018, Sherratt et al., 2023]. For instance, Johansson et al. (2019) [Johansson et al., 2019] found that ensembles of dengue forecasting models tended to be better calibrated than individual models. Dengue fever, a mosquito-borne viral disease, is a major public health challenge in tropical and subtropical regions worldwide. Previous studies have associated dengue outbreak risk with levels of urbanization, connectivity, and temperature suitability [Lee et al., 2021]. Brazil, in particular, has witnessed a dramatic increase in dengue incidence in recent years, with the disease spreading to new geographical areas [Codeco et al., 2022]. In 2024, the number of cases surpassed the sum of the preceding decade [Gurgel-Gonçalves et al., 2024]. Dengue expansion and intensified transmission may strain public health systems, potentially resulting in an increase in the number of related-deaths and hospitalizations. In anticipation of the 2025 dengue season, in June 2024, the Infodengue-Mosqlimate consortium launched a dengue forecast sprint called Brazilian Dengue 2024 Challenge (IMDC24) (Fig.1). Infodengue is a dengue early warning system that is active in Brazil since 2015, producing weekly reports for all municipalities (info.dengue.mat.br), while Mosqlimate (www.mosqlimate.org) is an open-source web-based platform designed to support the generation, comparison, and sharing of forecast models for dengue and other mosquito-borne diseases. The goal of the Sprint was to generate forecasts that could provide actionable insights, guiding resource allocation for effective prevention and control measures. To this end, we assembled a comprehensive public dataset that included case data, climatic, and demographic variables, with a custom API for easy data retrieval [Coelho et al., 2024]. To ensure alignment with the Brazilian health policy objectives, the forecasting targets were collaboratively established with the Ministry of Health (MoH). In addition, results needed to be delivered in a timely manner, facilitating evidence-based decision-making and strategic planning for the next epidemic season. This pioneering initiative in Brazil aimed at bridging the gap between research and real-world application, drawing inspiration from previous endeavors such as the 2015 CDC Dengue Forecasting Challenge [Johansson et al., 2019]. This paper describes the IMDC24 initiative, including its organization, implementation, and results. A total of six teams from four countries participated. The teams were invited to produce forecasts for two seasons (2023 and 2024), for five federative units (FU)(states), one for each of the five macroregions of Brazil. This comprised the validation phase, with a total of 10 predictions submitted by each team, which were scored using different methods (described in Section). At the end of the Dengue Forecast Sprint, the teams were invited to produce forecasts for 2025 (true forecasts). The submitted models were used to build two ensemble forecast models to predict the 2025 season.

**Figure 1:**
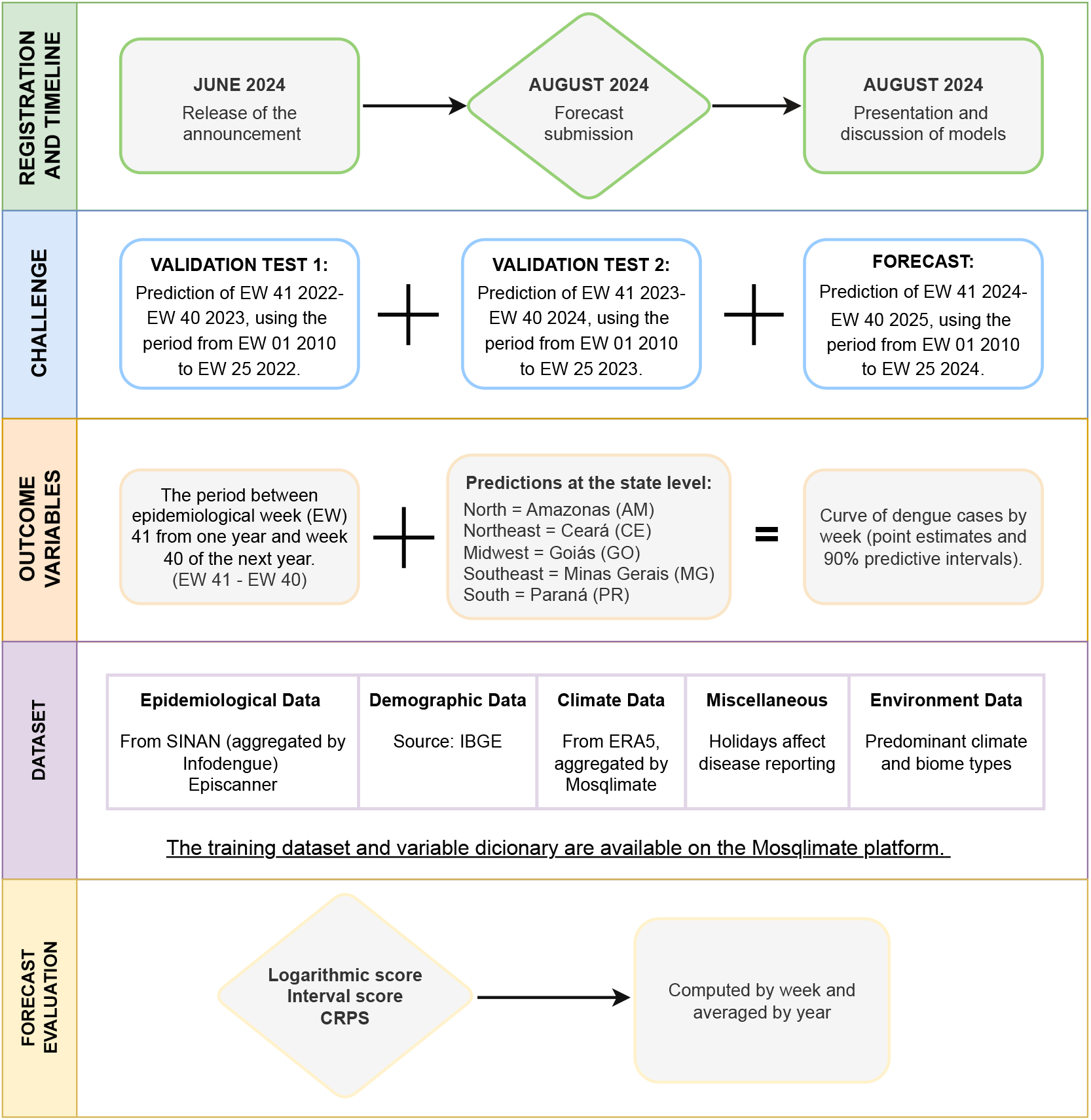
Diagram summarizing the Infodengue-Mosqlimate Dengue 2024 Challenge.

## Material and methods

The IMDC24 lasted approximately three months. The announcement was released on 20 June 2024, with a forecast submission deadline of 16 August 2024. The tight schedule was necessary to accommodate the needs of the decision-makers. A document describing the rationale, challenges, links to the data and rules was provided to all participants [Infodengue - Mosqlimate, 2024]. To participate, each team was required to fill out an online form, read and accept the instructions and rules and agree to use the training and validation as indicated. Then, a link to the sprint environment was provided.

The five Brazilian states included in the IMDC24 have unique climatic and socio-economic characteristics, urbanization levels, and dengue history (see Fig. S1). These states span a latitudinal gradient from the equatorial north in Amazonas (AM) to the subtropical south in Paraná (PR), with distinct seasonal temperature patterns. Additionally, the longitudinal range, from Goiás (GO) in the west to Minas Gerais (MG) and Ceará (CE), in the east, is marked by widely varying humidity levels. All selected states have undergone multiple dengue outbreaks in the last 20 years, with the 2024 season being the most severe in Minas Gerais, Paraná, and Goiás (see Fig. S1). Figure S1 delineates the two primary forecasting exercises proposed to the teams: the first challenge was to train models using data up to 2022 (white background) and test their forecasts against 2023 data (shaded blue), while the second challenge involved training models up to 2023 and testing them using data from the shaded green period (2024).

### Data

The IMDC24 dataset and its documentation were made available through the Mosqlimate platform [Coelho et al., 2024] and the interested reader is referred to Ganem et.al [Ganem et al., 2024], which describes how the mosqlimate platform works This comprehensive dataset included administrative, demographic, epidemiological, and climate data for all cities included in the challenge and additionaly for all 27 Brazilian states and 5570 municipalities.

#### Administrative and demographic data

Within each state, municipalities are organized into health districts. The names and codes of both administrative divisions were provided for each municipality. The estimated population size per municipality were obtained from the Brazilian Institute of Geography and Statistics - IBGE.

#### Epidemiological data and case definition

Case notification data of probable dengue cases at the date of symptom onset were made available aggregated by epidemiological week and municipality. A probable dengue case is confirmed through laboratory testing or clinical-epidemiological criteria, or is still under investigation. The Brazilian Ministry of Health uses this case definition for arboviruses surveillance. The dengue epidemiological year (here referred as epiyear) is defined as the period spanning from the epidemiological week (EW) 41 of a calendar year to the epidemiological week 40 of the following year. The dataset comprises 14 seasons of case notification data between epidemiological years 2010-2024 in addition to some derived measures, such as the effective reproductive number (*R*_*t*_) for dengue per epiweek and municipality, provided by Infodengue [Codeco et al., 2018]. A second group of descriptors came from the Episcanner tool [Araujo et al., 2024]. This tool fits Richard’s model to the dengue incidence data and estimates the basic reproduction number (*R*_0_), the peak week, the total outbreak size, and the outbreak duration. These metrics were available only for municipalities and years that had reported dengue transmission, defined as a minimum of 3 weeks with *R*_*t*_ *>* 1, and at least 50 cases reported since 2010.

#### Climate data

Meteorological variables were obtained from the ERA5 reanalysis product, encompassing temperature, pressure, humidity, and precipitation [Muñoz Sabater, 2019]. Hourly data were aggregated to epidemiological week for each municipality and the average, maximum, and minimum statistics were computed. The diurnal temperature range was also calculated and summarized in the same way. Furthermore, monthly aggregated climate anomaly indices - the El Niño Southern Oscillation (ENSO), the Indian Ocean Dipole (IOD), and the Pacific Decadal Oscillation (PDO) - were obtained from the National Oceanic and Atmospheric Administration (NOAA) (https://psl.noaa.gov/enso/mei/).

#### Other data

A set of variables fixed in time were also provided for each municipality: the predominant Koppen climate type, the predominant biome, and the average altitude. These variables were obtained from the Brazilian Institute of Geography and Statistics - IBGE.

Participants were allowed to use additional data as long as those data were made available to all participants. A description of all data is available in the supplementary material, section 4, Sprint data and parameters description.

### Challenges

A series of forecasting challenges were presented to the teams. The first two exercises utilized historical data (see details below) and served as benchmarks for validation and comparative analysis of predictions. These are depicted as shaded regions in Fig. S1. Then, teams were invited to provide forecasts for 2025 (i.e., forward-looking forecasts). For each challenge, the teams were instructed to provide the predictions for each of the selected states, compute point estimates and the (90%) uncertainty intervals, and upload them to the Mosqlimate platform.

#### First challenge

predict the weekly number of dengue cases for each selected state in the 2023 epiyear [EW 41/2022- EW 40/2023], using data covering the period from EW 1/2010 to EW 25/2022. Overall, the observed dengue incidence during this epiyear was not as high as in 2024, thus more comparable with historical levels.

#### Second challenge

predict the weekly number of dengue cases for each selected state in the 2024 epiyear [EW 41 2023- EW 40 2024], using data covering the period from EW 1/2010 to EW 25/2023. Dengue incidence in this epiyear was extremely high, posing a challenge for the predictive models.

#### Third challenge

The teams were tasked with generating forecasts for the 2025 epiyear using the same models employed for the first two targets. The forecast window was set to be from EW 41/2024 to EW 40/2025. As these targets pertain to future events, they are not subject to validation within this study.

### Parametric approximation and scoring

As previously mentioned, the predictions were presented as point estimates accompanied by uncertainty intervals. To aid tractability, we decided to apply a parametric approximation to the predictive distribution by using a log-normal distribution, which provided a good approximation of the median and the upper prediction interval – we give a detailed description of the fitting process in the supplementary material.

The performance of the models was evaluated using three scores: logarithmic score, continuous ranked probability score (CRPS), and interval score, available in the Python *scoringrules* package [Zanetta and Allen, 2024]. The scores for each model were computed by state, EW and averaged by epiyear.

The CRPS for a log-normal distribution was computed using the expression in Equation (1):

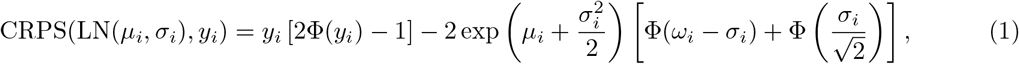

where Φ is the cumulative distribution function (CDF) of the standard normal distribution and 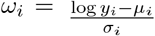, in which *y*_*i*_ refers to the cases observed in week *i, µ*_*i*_ is the forecast for week *i* and *σ*_*i*_ is the standard deviation of the forecast in week *i*.

The logarithmic score was computed as:

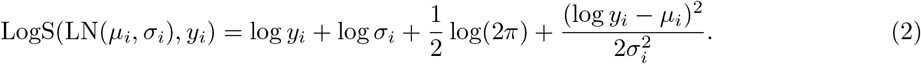

The interval score was calculated using the equation 3:

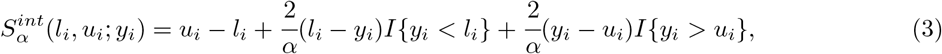

where *I* is the indicator function, *α* is the (significance) level of the interval, *u*_*i*_ the upper value of the interval at week *i*, and *l*_*i*_ the lower value.

### Ensemble models construction

Combining predictive models into ensembles is a well-established technique to improve forecast accuracy [Breiman, 1996, Gneiting and Raftery, 2005, Reich et al., 2019b]. To construct an ensemble from all the submitted models, we adopted a methodology based on the logarithmic pooling of the predictive probability distributions for each model [Carvalho et al., 2023] (see Section 2 of supplementary material for details). Two ensemble models (*E*_1_ and *E*_2_) were constructed: the first (*E*_1_) used equal weights and a linear mixture of predictive distributions as a baseline, while the second (*E*_2_) had its weights optimized to minimize the CRPS score of a logarithmic pooling of the ensemble predictions (see Fig. S6).

To assess the performance of the ensemble *e* models in comparison with each individual model *m*, we computed a skill score based on their CRPS score, defined as

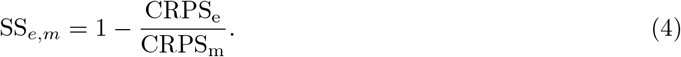

The 2025 forecasts, uploaded to the Mosqlimate platform, will be scored using the same methodology described in this study once the data become available at the end of the 2025 dengue epiyear in Brazil.

## Results

Six teams submitted probabilistic forecasts for the five Brazilian states. One of them submitted two models, totaling seven models (*M*_1_-*M*_7_) 1. The specific structures of each model and how they used the available datasets are described in Section 1 of supplementary material. Model types ranged from classical regression models to deep neural networks, with very diverse approaches.

**Table 1:**
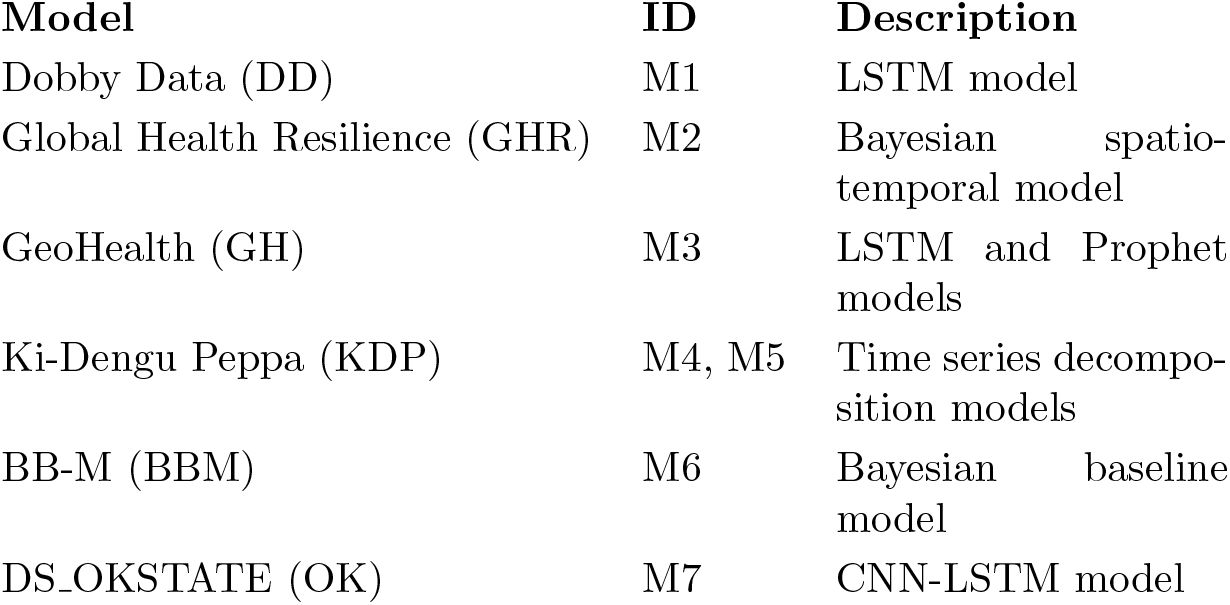
Submitted Forecast Models.

### Individual models performance

Overall, forecast accuracy and precision varied considerably among models for the 2023 and 2024 epiyears. Figures 2 show the forecasts for AM (a northern state with fewer cases and more endemic-like behavior) and MG (a southeastern state that experienced larger seasonal peaks in the previous year). Forecasts for the other states (CE, GO, and PR) are shown in Figs. S2–S4. Figure 3A displays the CRPS values for each state and model in 2023 and 2024. Interval scores are provided in Fig. S7, and logarithmic scores in Fig. S8. In Amazonas (AM), where the 2023 dengue epiyear was below average, two models overestimated the epidemic curve (*M*_4_ and *M*_7_), while the remaining models aligned well with the observed data. Among them, *M*_1_ and *M*_6_ were the most accurate based on the CRPS and the interval scores (Fig. 3A and S7).Based on the logarithmic score, the best models are *M*_6_ and *M*_2_ (Fig. S8). In 2024, during a period of above-average seasonal incidence, models *M*_2_ and *M*_7_ exhibited the best performance based on the CRPS score, while models *M*_3_, *M*_6_, and *M*_7_ were optimal according to the interval score. As observed in 2023, models *M*_2_ and *M*_6_ continue to outperform others based on the logarithmic score.

**Figure 2:**
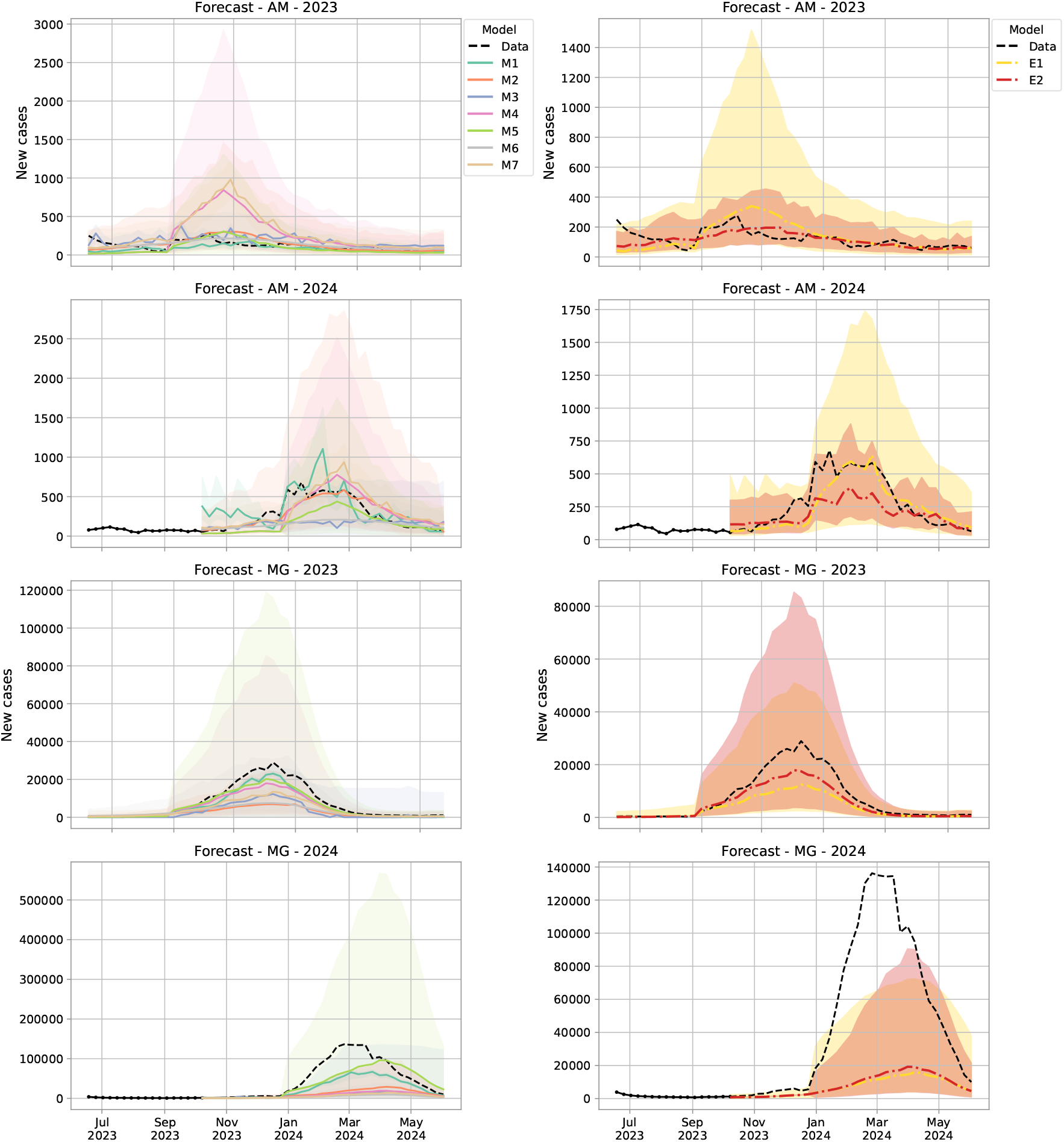
Forecasted (median and 90% uncertainty intervals) versus observed dengue cases by week for two selected Brazilian states (Amazonas - AM and Minas Gerais - MG) and epiyears 2023 and 2024 produced by individual models (M1-M7) (first column) and ensemble methodologies (E1 and E2) (second column).

**Figure 3:**
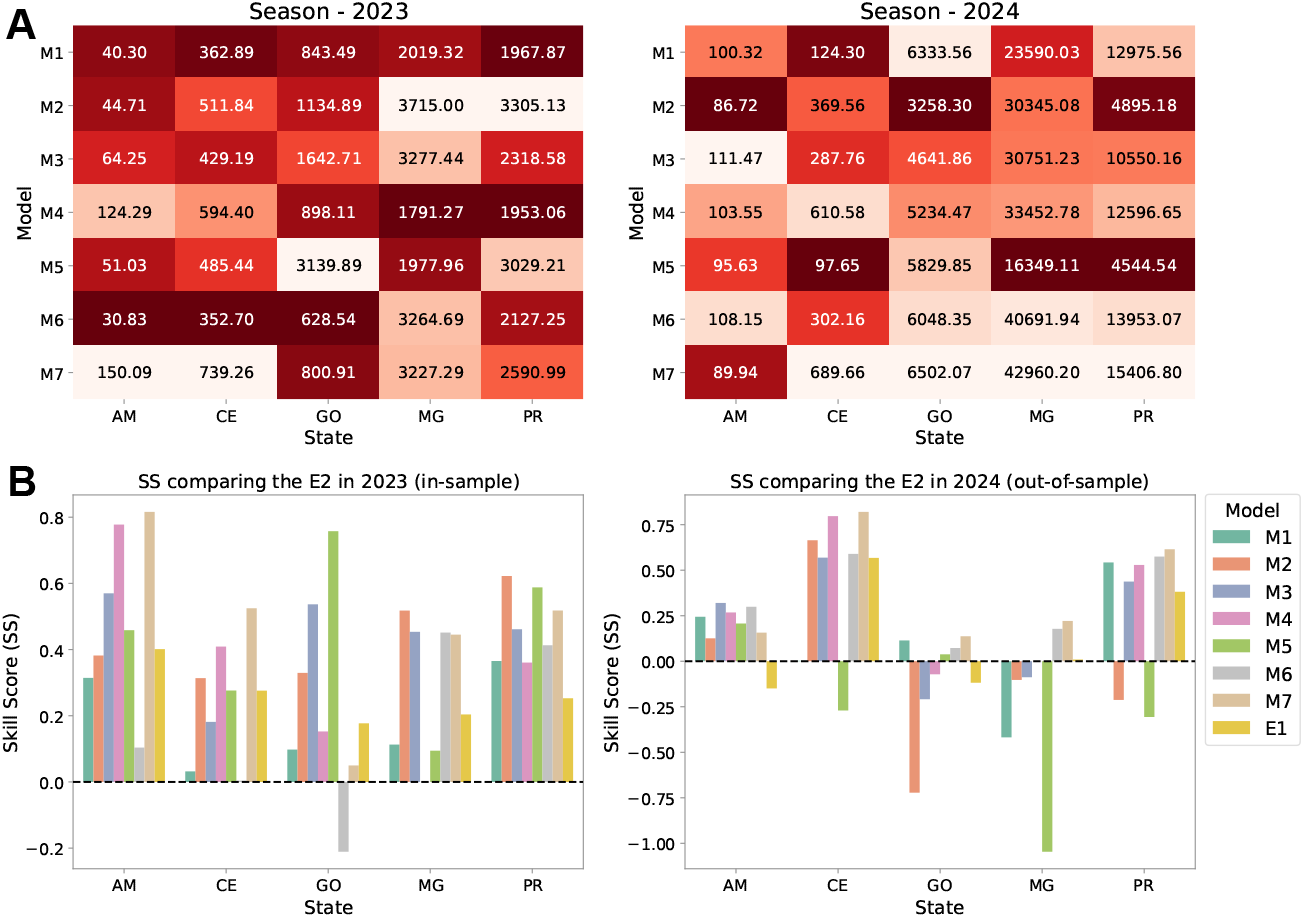
Forecasting models performance based on the continuous ranked probability score (CRPS) and the Skill Score (SS). (A) CRPS values for the individual models by state for epiyears 2023 and 2024, with lower values indicating better performance. The CRPS values represent the average score over the entire prediction window. (B) Skill Score (SS) comparing the ensemble model *E2* with the individual models and the ensemble model *E1* by state for 2023 (in-sample) and 2024 (out-of-sample). Positive *SS* values are interpreted as superior predictive skill of the ensemble model *E2*.

In Minas Gerais (MG), both years were characterized by above-average dengue activity (Fig. 2). In 2023, *M*_4_ had the best CRPS scores (Fig. 3A), but not the best interval and logarithmic scores (Fig. S7 and S8, followed by *M*_5_ and *M*_1_). In 2024, none of the models predicted the earlier onset of the epidemic curve. The models tended to underestimate the cases, with *M*_5_, *M*_3_ and *M*_2_ performing best according to the CRPS, interval score, and logarithmic score respectively.

Ceará (CE) experienced below-average dengue epiyears in 2023 and 2024 (Fig. S2). In 2023, all models tended to overestimate the epidemic curve, while in 2024, *M*_1_ and *M*_5_ aligned well with the data, with considerably better interval scores than the other ones (Fig. S7). According to the logarithmic score, models *M*_6_ and *M*_2_ demonstrated the best overall performance (Fig. S8).

For Goiás (GO), all models tended to overestimate the epidemic curve in 2023, a below-average epiyear (Fig. S3). The estimates were consistent across models, except *M*_5_, which exhibited poorer performance. The 2024 epiyear in this state was exceptionally high and no model successfully captured it. Only two models (*M*_2_ and *M*_3_) generated uncertainty intervals that encompassed the observed level.

In Paraná (PR), the 2023 epiyear was moderate while 2024 was the highest epiyear in the study period. While the best models were *M*_1_ and *M*_4_, in 2023, according to the CRPS scores, they greatly underestimated the peak (Fig. S4 and 3A). Meanwhile, *M*_4_ and *M*_6_ were the best according to the interval score (Fig. S7), while *M*_6_ and *M*_2_ were top-ranked according to the logarithmic score. For 2024, all models underestimated the severity of the 2024 epiyear, with *M*_5_ and *M*_2_ as the best models according to the CRPS score (Fig. 3), while *M*_3_ and *M*_2_ were the top performers on the other two score methods (Fig. S7 and S8).

We report the weekly values of the scores in section 3.2 of the supplementary material. In general, scores were worse when the incidence was higher. According to the CRPS score (Fig. 3A), the best-performing models in 2023 were *M*_6_ and *M*_4_, while in 2024, *M*_2_ and *M*_2_ demonstrated the highest performance.

### The ensemble models

The second column of Fig. 2 shows the forecasts produced by the two ensemble models, considering an implementation with equal weights (*E*_1_) and the logarithmic pool with CRPS informed weights (*E*_2_), for the states of AM and MG. The forecasts for the remaining states are shown in Figures S2, S3, and S4. Fig. 3B shows the results of the skill score of the *E*_2_ model compared to *E*_1_ and the individual models (*M*_1_ - *M*_7_). The ensemble model *E*_2_ outperformed *E*_1_ and most individual models in 2023 (in-sample forecasts), except *M*_6_ for GO. For the out-of-sample forecasts, the results were mixed. The *E*_2_ model outperformed most models for AM, CE and PR. However, for GO and MG, the *E*_2_ ensemble model did not show a clear improvement over individual models while predicting the highly atypical seasons of 2024.

A notable contribution of the ensemble models is the reduction of the forecast uncertainty. This is clear for all target states.

### Forecasting the 2025 dengue epiyear

Fig. 4 shows, for each of the five states, two forecast curves for the 2025 epiyear. The curves correspond to the ensembles computed using the (*E*_2_) methodology, based on the logarithmic pooling of the predictions. The ensemble *E*_2_(2023), shown in green, use the pooling weights to minimize the CRPS of the models in the 2023 epiyear, while the ensemble *E*_2_(2024), in blue, does it in the 2024 epiyear. For all states, the 2024-trained models produced higher epidemic curves, possibly capturing the expectation of a worse season after a sequence of increasing waves.

**Figure 4:**
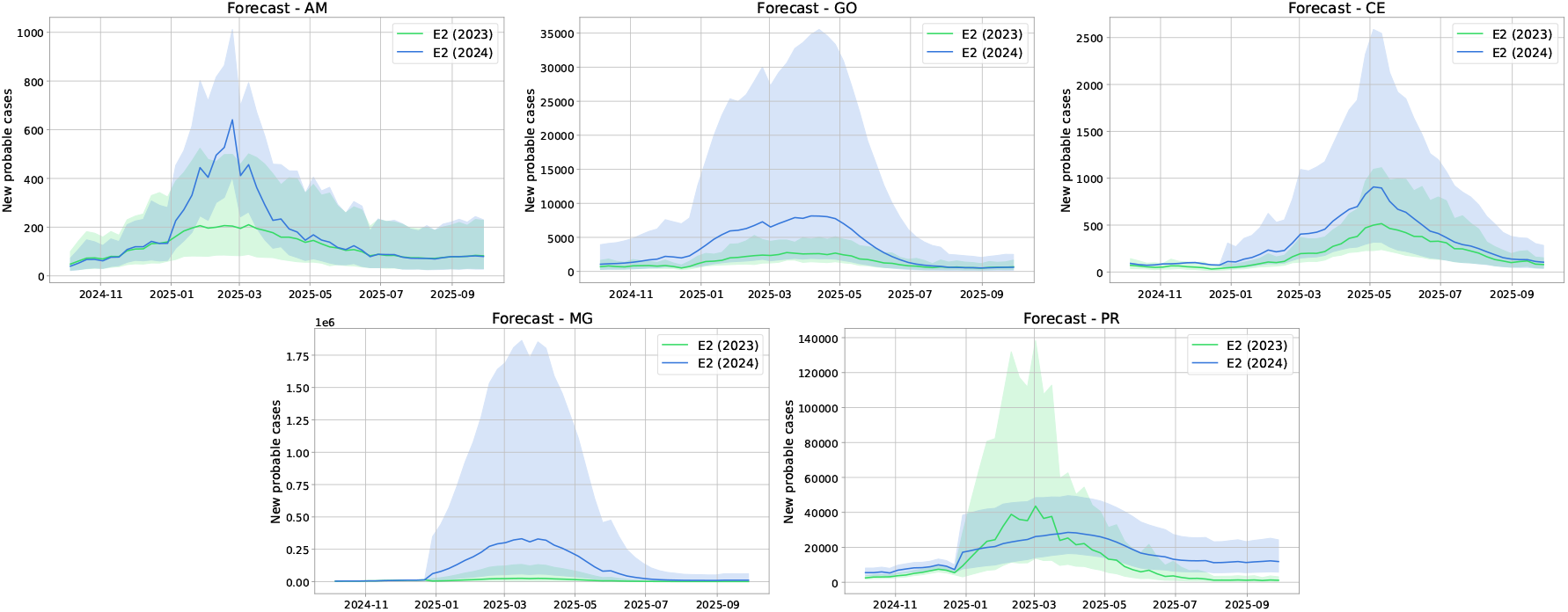
Forecasting of weekly probable dengue cases (median and 90% uncertainty interval) for the 2025 epiyear across the five Brazilian states included in the IMDC24. The green line represents the forecasted values derived from the ensemble model based on 2023 predictions, while the blue line represents the forecasted values derived from the ensemble model based on 2024 predictions.

## Discussion and directions for future research

Planning for the upcoming dengue season is a significant challenge for health officials in endemic countries. Action and contingency plans are updated at local and national levels, requiring precise estimates of disease burden and accurate epidemic curves to ensure effective resource allocation for prevention, control, and primary care interventions.

This study demonstrates that organizing a multi-model sprint for action is feasible and can deliver timely results, provided certain conditions are met: access to comprehensive and harmonized data, robust infrastructure for model submission and evaluation, and a well-established community of modelers. The full results of the Challenge were published in September as a technical report (in Portuguese) addressed to the Brazilian Ministry of Health, ensuring it reached key decision-makers in the country [Codeço Coelho et al., 2024]. Additionally, several webinars were organized to elucidate and discuss the results. Online attendance reached almost a thousand people, including a large number of public health professionals, hailing from different Brazilian cities.

No single model consistently excelled across all forecast targets, largely due to the long lead times proposed for the forecasts, which posed a substantial challenge – see [Johansson et al., 2019]. While most models performed well for relatively typical seasons, neither individual models nor ensemble models provided accurate results for the extreme epidemic during the 2024 epiyear. This season was atypically severe, a phenomenon that could be partially attributed to climate change, which has altered environmental conditions, expanding dengue’s reach to previously unaffected areas. Another potential factor that hampered models in 2024 is the increased Oropouche fever incidence, which presents symptoms similar to dengue [Barçante and Cherem, 2025] and could have been reported as dengue cases. Addressing these uncertainties requires enhanced laboratory confirmation, although this poses a challenge in a country as large as Brazil. The performance of the ensemble models could be enhanced by incorporating additional models and extending the number of epiyears used for weight calculation. In this study, weights were derived solely from data from the year 2023 and subsequently applied to 2024.

Overall, models *M*_1_ and *M*_6_ were among the top performers across states in the 2023 epiyear, while in the 2024 epiyear, models *M*_2_ and *M*_5_ tended to rank higher according to the CRPS score, as shown in Fig. 3A. Notably, the models *M*_2_, *M*_5_ and *M*_6_ are Bayesian, underscoring the advantages of this methodology for generating probabilistic forecasts compared to machine learning and deep learning models.

This study employed three different scoring methods to evaluate models, which did not always agree. Scoring remains an important avenue for future development, in particular concerning the development of scores that capture the key aspects involved in public health decision-making [Bosse et al., 2022].

Some choices made for the challenge posed constraints for the extrapolation of results to other settings. For instance, selecting states with a history of high dengue endemicity facilitated model development due to the availability of long time series and outbreak sequences for training. However, we did not assess model performance in areas with low endemicity or recent disease emergence – as seen, for instance, in Brazil’s southernmost states – thus neglecting the important epidemiological task of predicting potential outbreaks or an unprecedented increase in historically low areas.

The models presented here do not encompass all possibilities for forecasting dengue. For example, they did not integrate climate forecasts, or virological surveillance data, which could enhance predictability. As we write this paper at the start of 2025, the forecasts have yet to be tested, and outcomes will be displayed on the IMDC website (sprint.mosqlimate.org). Thus far, the ensemble models appear to have overestimated the true incidence curve.

## Conclusion

The IMDC24 initiative underscores that planning for future epidemics necessitates multi-model approaches to account for epistemic and probabilistic uncertainties. The quality of models can be enhanced by robust surveillance systems, high data quality, and a deep understanding of causal links. We conclude that forecast scenarios are vital tools in the public health toolkit, providing an additional piece of epidemiological intelligence alongside other decision-support tools. Models should be viewed as scenarios for action, built on incomplete knowledge, reinforcing, as a recommendation, the importance of joint work, of engagement with health professionals who work in surveillance.

## Acknowledgements

FC acknowledges support from the Wellcome Trust (Mosqlimate 218987/Z/19/Z). RL and CTC acknowledge support from the Wellcome Trust (HARMONIZE 224694/Z/21/Z and IDExtremes 226069/Z/22/Z). RL acknowledges EU’s Horizon Europe research and innovation programme (E4Warning; grant agreement 101086640 and IDAlert; grant agreement 101057554) and a Royal Society Dorothy Hodgkin Fellowship. LPF and LSB are supported by a grant from the Inova/Fiocruz/Oswaldo Cruz Foundation and the Department of Public Health Emergencies of the Secretariat for Health and Environmental Surveillance of the Ministry of Health (DEMSP/SVSA/MS) - Brazil [VPPCB-002-FIO-20-2-27], and by the National Council for Scientific and Technological Development (CNPq) and the Department of Science and Technology of Secretariat of Science, Technology, Innovation and Health Complex of the Ministry of Health of Brazil (Decit/SECTICS/MS) - Brazil [444896/2023-6]. LSB acknowledges support from CNPq – Brazil [310530/2021-0] and FAPERJ – Brazil [E-26/201.277/2021 and E-26/204.098/2024], RML acknowledges the HORIZON-MSCA-2022-PF-01 grant (project number 101109642), PM acknowledges support from The Letten Prize (https://lettenprize.com/).

## Data availability

The data underlying this article are available for download from Zenodo [Coelho et al., 2024] as well as on the mosqlimate platform from where it can be freely downloaded through its API.

