## Supplementary material for "Leveraging probabilistic forecasts for dengue preparedness and control: the 2024 Dengue Forecasting Sprint in Brazil"

### Supplementary material for Leveraging probabilistic forecasts for dengue surveillance: the 2024 Mosqlimate-Infodengue Sprint

May 12, 2025

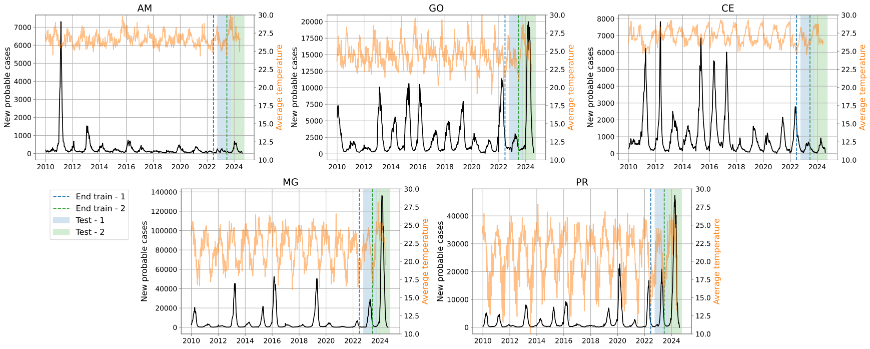


**Fig. S1.** Time series of probable dengue cases (black curve) and average temperature (orange curve) across the five target states from 2010 to 2024. The dashed vertical lines indicate the end of the training periods. The shaded regions denote the testing periods. The blue-shaded region corresponds to the first challenge: training up to week 25 of 2022 and forecasting from weeks 41 of 2022 to 40 of 2023. The green-shaded region corresponds to the second challenge: training up to week 25 of 2023 and forecasting from weeks 41 of 2023 to 23 of 2024.

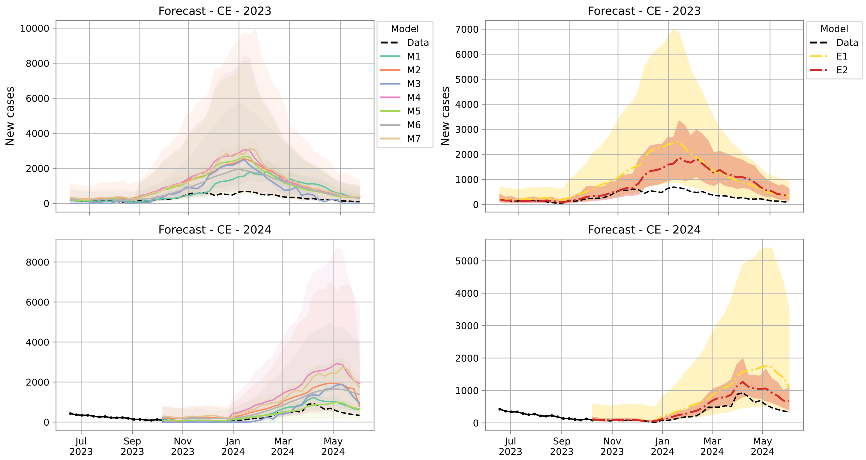


**Fig. S2.** The first column of the figures displays the forecasted number of probable dengue cases for CE in 2023 and 2024. The acronyms M1 to M7 represent the forecast generated by the individual models. The second column of the figures presents the forecasted dengue cases for CE in 2023 and 2024 using ensemble methodologies. The forecasts labeled E1 and E2 correspond to the two ensemble approaches.

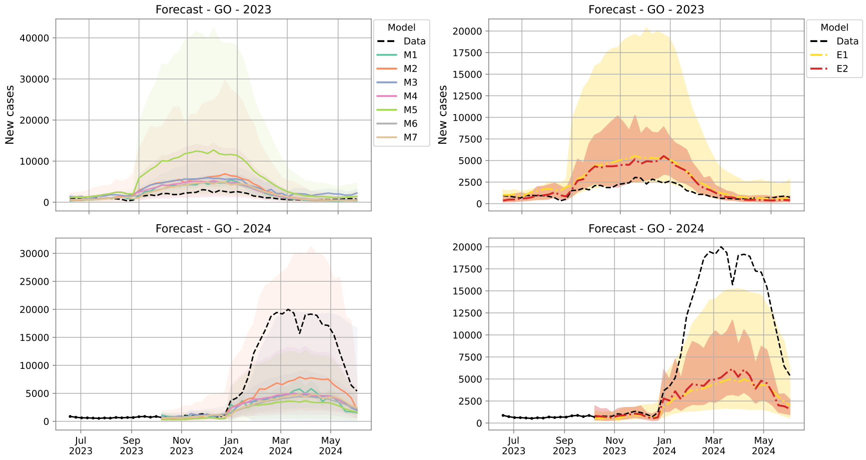


**Fig. S3.** The first column of the figures displays the forecasted number of probable dengue cases for GO in 2023 and 2024. The acronyms M1 to M7 represent the forecast generated by the individual models. The second column of the figures presents the forecasted dengue cases for GO in 2023 and 2024 using ensemble methodologies. The forecasts labeled E1 and E2 correspond to the two ensemble approaches.

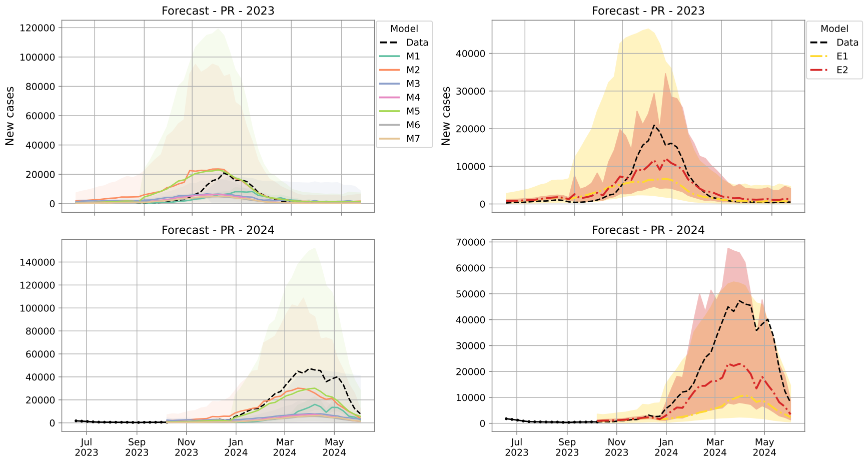


**Fig. S4.** The first column of the figures displays the forecasted number of probable dengue cases for PR in 2023 and 2024. The acronyms M1 to M7 represent the forecast generated by the individual models. The second column of the figures presents the forecasted dengue cases for PR in 2023 and 2024 using ensemble methodologies. The forecasts labeled E1 and E2 correspond to the two ensemble approaches.

### 1 Submitted forecast models

In this section, we briefly describe the models submitted to the sprint and included in the ensembles. They are labeled $M_1$ to $M_7$ models.

#### 1.1 $M_1$: Long Short-Term Memory (LSTM) model

$M_1$ is a deep neural network model based on LSTM layers from the *tensorflow package* Abadi et al. (2015). For each of the five states, models with different architectures were trained using data at their health region level. State-level predictions were obtained by adding the series from all health districts. The confidence intervals were computed using the dropout technique (Srivastava et al., 2014).

To predict the dengue season for year $y$, data from EW 41 of $y-3$ to EW 25 of $y-1$ were used. The input features included weekly case numbers, the ENSO (El Niño Southern Oscillation) index, epiweek, and the epidemiological parameters of each health district: 'R0', 'total_cases', and 'peak_week'. A feature called 'perc_geocode' was also included, which represents the fraction of cities, each year, in the health district, that reported dengue epidemics. An exception was made for 'PR' for which only case counts, ENSO, and epiweek were used. Case counts were normalized using the Box-Cox transformation.

For each state, three architectures were explored: 1. model with two concatenated LSTM layers; 2. model combining an attention layer with an LSTM layer;

3 model that concatenated the outputs from the first two architectures. The first architecture performed best for the states of PR, MG, and CE. The second architecture showed better performance in GO, while the third performed best in AM. We also compared the mean squared logarithmic error (MSLE) and mean squared error (MSE) as loss functions. Each LSTM layer consisted of 32 hidden units, and the models were trained with a batch size of four. The code is available at https://github.com/eduardocorrearaujo/lstm_transf_to_state.

#### 1.2 $M_2$: Bayesian spatio-temporal model

This is a Bayesian spatio-temporal model with a negative binomial likelihood. First, dengue cases were aggregated by health district $n = 450$) and time (weeks, $n = 52$). The model covariates consist of 3 meteorological variables, expressed at a monthly time scale: i) 3-month moving average of the mean temperature, ii) 12-month Standardized Precipitation Index (SPI), and iii) 1-month SPI. SPI represents the total precipitation in units of standard deviation from the historical average over a given period, assuming a gamma-fitted distribution. The model also includes a set of spatio-temporal random effects.

Let $Y_{t,h}$ be the number of dengue cases at time $t$ in health district $h$. The model is specified as follows:

$$Y_{t,h} \sim \text{NegBin}\big(\lambda_{t,h},\, \kappa\big), \tag{1}$$

where $\kappa > 0$ is the overdispersion parameter. The mean $\lambda_{t,h}$ is modeled as

$$\log\big(\lambda_{t,h}\big) = \upsilon_h + \phi_h + \gamma_{w[t],\,s[h]} + \delta_{y[t],\,m[h]} + \Big(\text{SPI}_{12,\text{lag5}} \times \text{SPI}_{1,\text{lag1}} \times \text{Tmean}_{3,\text{lag3}}\Big), \tag{2}$$

where

- $\upsilon_h$ is an iCAR (intrinsic conditional autoregressive) random effect for each health district $h$,
- $\phi_h$ is an i.i.d. Gaussian random effect for each health district,
- $\gamma_{w[t],\,s[h]}$ is a second-order random walk (RW2) effect by week $w[t]$ and state $s[h]$,
- $\delta_{y[t],\,m[h]}$ is a first-order random walk (RW1) effect by year $y[t]$ and macroregion $m[h]$,
- $\text{SPI}_{12,\text{lag5}}$ is the 12-month SPI, measuring long-term precipitation levels, lagged by 5 months
- $\text{SPI}_{1,\text{lag1}}$ is the 1-month SPI, measuring short-term precipitation levels, lagged by 1 months
- $\text{Tmean}_3$ is the 3-month moving average of the temperature, lagged by 3 months

- The product $\text{SPI}_{12,\text{lag5}} \times \text{SPI}_{1,\text{lag1}} \times \text{Tmean}_{3,\text{lag3}}$ specifies the full factorial interaction among $\text{SPI}_{1,\text{lag1}}$, $\text{SPI}_{12,\text{lag5}}$, and $\text{Tmean}_3$ comprising the three main effects, the three two-way interactions ($\text{SPI}_{1,\text{lag1}}$:$\text{SPI}_{12,\text{lag5}}$, $\text{SPI}_{1,\text{lag1}}$:$\text{Tmean}_3$, and $\text{SPI}_{12,\text{lag5}}$:$\text{Tmean}_3$), and the three-way interaction ($\text{SPI}_{1,\text{lag1}}$:$\text{SPI}_{12,\text{lag5}}$:$\text{Tmean}_3$) for a total of seven terms.

Since the model includes 3 climatic variables evaluated at 3 different lags, the predictions for 2025 dengue season were based on climate forecasts. Specifically, climate forecasts with 1 to 7 month lead time issued in August 2024 were used to produce predictions. For months where forecast was not available we used climatological means.

The model was fitted using the `R-INLA` software (Lindgren and Rue, 2015; Rue et al., 2009a). Samples from the posterior predictive distribution were used to generate forecasts for the following season at the state level by appropriate aggregation. All code is available at https://github.com/giovemoiran/infodengue-sprint-lsl.

##### 1.3 $M_3$: Long Short-Term Memory (LSTM) Model and Prophet model

Initially, to improve model performance and reduce redundancy in the dataset, we applied variance threshold selection (Guyon and Elisseeff, 2003) to remove low-variance climate features, ensuring that only informative variables were retained. The remaining features, which included various climate indicators such as temperature, precipitation, and humidity, were further processed using Principal Component Analysis (PCA) (Jolliffe, 2002), preserving 95% of the total variance. The transformed dataset was then used as input for both models.

**LSTM.** LSTM (Greff et al., 2016; Hochreiter and Schmidhuber, 1997) is a type of recurrent neural network (RNN) specifically designed to handle long-term dependencies in time series data. The LSTM model was trained to forecast dengue cases based on historical dengue incidence and climate variables. A standard LSTM unit consists of three key gates:

- Forget Gate: Determines which information should be discarded.

$$f_t = \sigma(W_f \cdot [h_{t-1}, x_t] + b_f)$$

- Input Gate: Updates the cell state with new information.

$$i_t = \sigma(W_i \cdot [h_{t-1}, x_t] + b_i)$$
$$\tilde{C}_t = \tanh(W_C \cdot [h_{t-1}, x_t] + b_C)$$

- Output Gate: Determines the final output of the unit.

$$o_t = \sigma(W_o \cdot [h_{t-1}, x_t] + b_o)$$
$$h_t = o_t \cdot \tanh(C_t)$$

The optimal LSTM model for each state and task was determined through grid search, selecting the model with the lowest Mean Absolute Percentage Error (MAPE).

**Prophet Model.** Prophet (Taylor and Letham, 2018) is an additive regression model designed for time series forecasting. It decomposes time series data into three components: trend, seasonality, and holiday effects:

$$y(t) = g(t) + s(t) + h(t) + \epsilon_t$$

where $g(t)$ represents the trend component, modeled using a piecewise linear or logistic growth function; $s(t)$ captures seasonal variations using Fourier series; $h(t)$ incorporates the effect of holidays or external regressors; $\epsilon_t$ is the error term.

For dengue forecasting, Prophet was configured to capture yearly and weekly seasonality while incorporating climate variables as external regressors.

Model selection was performed using MAPE to determine whether LSTM or Prophet provided the best predictions for each state (Chen and Moraga, 2024). The code is available at `https://github.com/ChenXiang1998/Infodengue-Sprint/tree/main/model`.

##### 1.4 $M_4$ and $M_5$: Time series decomposition models

###### $M_4$ Weekly and yearly (iid) components

$M_4$ is based on a structural decomposition designed to model counting series employing a Poisson distribution framework. Within this distribution, the log intensity undergoes variations over time, defined by the sum of weekly and yearly components. Using a Poisson process structure with stochastic intensity implies that the process is characterized by a Cox process. The weekly component is defined through a first-order autoregressive process, while the yearly component is assumed to be independently and identically distributed. The proposed model is expressed as follows:

$$\begin{aligned}
Y_t &\sim \text{Poisson}(\exp(\lambda_t)), \\
\lambda_t &= \gamma_t(\text{week}) + \delta_t(\text{year}), \\
\gamma_t(\text{week}) &= \beta\gamma_{t-1}(\text{week}) + \eta_\gamma, \quad \text{week} = 1, \ldots, 52, \\
\delta_t(\text{year}) &\overset{\text{iid}}{\sim} N(0, \sigma^2_{\eta_\delta}), \quad \text{year} = 2010, \ldots, 2023.
\end{aligned}$$

The resulting Bayesian hierarchical structure allows us to perform inference procedures within the INLA framework, which provides accurate and efficient approximations on Bayesian hierarchical models that can be represented as latent Gaussian models.

##### $M_5$: Weekly and yearly (RW1) components

$M_5$ is similar to $M_4$ but assumes the yearly component as a random walk while the weekly component remains as a first-order autoregressive process.

$$\begin{aligned} Y_t &\sim \text{Poisson}(\exp(\lambda_t)), \\ \lambda_t &= \gamma_t(\text{week}) + \delta_t(\text{year}), \\ \gamma_t(\text{week}) &= \beta\gamma_{t-1}(\text{week}) + \eta_\gamma, \quad \text{week} = 1, \ldots, 52, \\ \delta_t(\text{year}) &= \delta_{t-1}(\text{year}) + \eta_\delta, \quad \text{year} = 2010, \ldots, 2023. \end{aligned}$$

Both models were implemented in R (R Core Team, 2022) version 4.4.1 using the **R-INLA** package (Lindgren and Rue, 2015; Rue et al., 2009a), without the inclusion of covariates. The full code is available at `https://github.com/fernandacvalente/kidenguPeppa`.

#### 1.5 $M_6$: Bayesian baseline model

This is a Bayesian negative binomial model with week and season as random effects. Let $Y_{t,h}$ be the number of dengue cases at time $t$ and health district $h$. We opt to use data from 2015 and to avoid noisy data we aggregate cases in space (health region) and time (weeks). Hence, the model is the following:

$$Y_{t,h} \sim \text{NegBin}(\lambda_{t,h}, \phi_h), \tag{3}$$

where $\phi_h > 0$ is an overdispersion parameter and $\lambda_{t,h}$ is the expected number of dengue cases at time $t$ in health district $h$, modelled as

$$\log(\lambda_{t,h}) = \alpha_h + \beta_{w[t],h} + \gamma_{s[t],h}, \tag{4}$$

where $\alpha_h$ are health district-specific intercepts, $\beta_{w[t],h}$ cyclic second-order random walk effects by week $w$ and health district $h$, and $\gamma_{s[t],h}$ independent Gaussian random effects for season $s$ and health district $h$. A season is defined as a period starting from the 41st epidemiological week (epiweek) of a given year and ending at the epiweek 40 of the following year.

The model is then completed by the default priors in INLA and the samples from the posterior predictive distribution were used to calculate the prediction for the next season by federal unit using adequate aggregations (Rue et al., 2009b, 2017). The entire code is available at `https://github.com/lsbastos/bb-m`.

#### 1.6 $M_7$: Convolutional Neural Network - Long Short-Term Memory (CNN-LSTM) model

$M_7$ is a deep neural network model composed of CNN layers, followed by LSTM layers, with a dense layer added to the output. In other words, after the CNN layers, the extracted features are passed to the LSTM layers – an advanced, recurrent neural network (RNN) – which utilize *gates* as described by Hochreiter and Schmidhuber Hochreiter and Schmidhuber (1997). This hybrid architecture of CNN-LSTM is particularly effective for time-series forecasting, as it captures both short-term local trends and long-term dependencies, and was developed in the context of Mpox prediction Das (2024). The architecture is displayed in Fig S5 and detailed in Table S1.

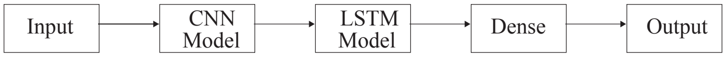


**Fig. S5.** Hybrid CNN-LSTM network architecture: The architecture of the CNN-LSTM model consists of two sub-models: the 1D CNN model (Conv1d), which processes sliding windows of the time series to extract local patterns and features, such as short-term trends, and the LSTM model, which captures long-term dependencies and relationships across different parts of the time series.

In this work, we used only epidemiological weekly (EW) of dengue cases in Brazilian states. Our forecasting method is described by Algorithm 1, which addresses three tasks: validation test 1, validation test 2, and forecast. The first task, validation test 1, focuses on predicting the weekly number of dengue cases by state for the *2022-2023 season* $[EW41\ 2022 - EW40\ 2023]$, utilizing historical data from *EW 01 2010* to *EW 25 2022*. The second task, *validation test 2*, predicts dengue cases for *2023-2024 season* from EW 41 2023 to EW 40 2024, based on data from *EW 01 2010* to *EW 25 2023*. The final task, the *forecast*, aims to predict the weekly number of dengue cases for the 2024-2025 season $[EW41\ 2024 - EW40\ 2025]$, using data from *EW 01 2010* to *EW 25 2024*.

**Supplementary Table S1.** CNN-LSTM Model Architecture and forecast

| Component | Description |
|---|---|
| Purpose | Defines and compiles a hybrid CNN-LSTM model for time series forecasting. |
| Inputs | $(n_{\text{steps}}, n_{\text{features}})$ : takes time series input data<br>$n_{\text{steps}}$ : number of time steps in input sequence<br>filters : number of filters in Conv1D layers<br>kernel_size : size of convolutional kernel<br>learning_rate : 0.0001<br>filters_vec : [32, 64, 128, 256, 512]<br>activation : ReLU |
| Architecture | - Conv1D(filters, kernel_size, activation = 'relu') (feature extraction)<br>- Conv1D(filters//2, kernel_size, activation = 'relu') (further feature extraction)<br>- MaxPooling1D(pool_size =1) (dimensionality reduction)<br>- LSTM(50, activation='relu') (captures temporal dependencies)<br>- Dense(30) (intermediate layer)<br>- Dense(10) (intermediate layer)<br>- Dense(1) (final output layer) |
| Optimizer and Loss | Adam optimizer, Mean Squared Error (MSE) loss function. |
| Output | Compiled CNN-LSTM model ready for training and evaluation. |
| Model forecast and Ensemble | Filters in filters_vec produce the forecasts along with the RMSE scores $R_i$. Using the RMSE scores $R_i$ from $M_i$ models, we define the penalty for each model as: $\hat{w}_i = \frac{1}{R_i}$. These penalties are then normalized to obtain the final weights: $\omega_i = \frac{\hat{w}_i}{\sum_{i=1}^{M} \hat{w}_i}$. The final ensemble prediction is $y_{\text{ensemble}} = \sum_{i=1}^{\#\text{ of } M_i} \omega_i y_i$, where $y_i$ represents the prediction vector from the $i$-th model. |

A key step in our algorithm is employing the expanding window training approach, progressively shifting the forecasting window while integrating forecasting residuals into training. The code can be found at https://github.com/haridas-das/DS_OKSTATE.

**Algorithm 1** Hierarchical model training and forecasting with expanding window

**Require:** Historical data $\mathcal{D}$ partitioned by epidemiological weeks (EW) with the levels $L \in \{0, 1, \ldots, 14\}$.

**Ensure:** Forecasts for all levels and update the residual feature.

**Step 0: Initialization**

Initialize storage for residuals and predictions:

$$\begin{aligned} \text{residuals} &\leftarrow \mathbf{0} \quad \text{(zero vector of size 40, EW 01, 2010 to EW 40, 2010)} \\ \text{predictions} &\leftarrow [] \quad \text{(empty list for storing predictions)} \end{aligned}$$

**for** level $i = 0$ to $L$ **do**

**Step 1: Define training, test, and prediction windows**

$$\begin{aligned} X_{\text{train}}, y_{\text{train}} &\leftarrow \text{EW01, 2010 to EW25, (2010+i)} \\ X_{\text{test}}, y_{\text{test}} &\leftarrow \text{EW40, (2010+i) to EW39, (2011+i)} \\ X_{\text{pred}}, y_{\text{pred}} &\leftarrow \text{EW41, (2010+i) to EW40, (2011+i)} \end{aligned}$$

**Step 2: Add residuals as an additional feature to training data**

$$X_{\text{train}} \leftarrow X_{\text{train}} \cup \text{residuals}[i]$$

**Step 3: Model training**

Train the model $\mathcal{M}$ using $X_{\text{train}}$ and predict on the test and forecasting window:

$$\begin{aligned} \hat{Y}_{\text{test}} &\leftarrow \mathcal{M}(X_{\text{test}}) \\ \hat{Y}_{\text{pred}} &\leftarrow \mathcal{M}(X_{\text{pred}}) \end{aligned}$$

**Step 4: Compute and update residuals**

$$\begin{aligned} &\text{Residuals}[i] \leftarrow y_{\text{pred}} - \hat{Y}_{\text{pred}} \\ &X_{\text{train}} \leftarrow X_{\text{train}} \cup \text{residuals}[i] \text{ for EW40, (2010+i) to EW25, (2010+i)} \end{aligned}$$

**Step 5: Fake test and fake pred data (for levels $L - 2$ to $L$)**

**if** $i \geq L - 2$ **then**

$$\begin{aligned} &X_{\text{pred}} \leftarrow \text{Average of } X_{\text{pred}} \text{ from levels 0 to } (i-1) \\ &y_{\text{pred}} \leftarrow \text{Average of } y_{\text{pred}} \text{ from levels 0 to } (i-1) \\ &X_{\text{test}} \leftarrow X_{\text{pred}} \\ &y_{\text{test}} \leftarrow y_{\text{pred}} \end{aligned}$$

Repeat Steps 3 and 4 with fake data.

**end if**

**Step 6: Predict and store the prediction for the level**

$$\begin{aligned} \hat{Y}_{\text{prediction}} &\leftarrow 10 \mathcal{M}(X_{\text{prediction}}) \\ \text{Predictions}[i] &\leftarrow \hat{Y}_{\text{prediction}} \end{aligned}$$

**end for**

**Step 7: Return all predictions**

Return predictions for all levels.

### 2 Ensemble models

#### 2.1 Dynamic ensemble *via* logarithmic pooling

Combining probabilistic forecasts into an ensemble is an efficient way to gather information from all models and build more accurate forecasts than can be achieved using any individual model. Since our predictive distributions of dengue cases were represented by unimodal distribution, we employed a model combination procedure that preserved unimodality, thus facilitating interpretation by decision-makers. To achieve this, we combined the approximate predictive distributions (see below) from all models using logarithmic pooling (Carvalho et al., 2023; Genest et al., 1984).

Let $f_1, \ldots, f_K$ be predictive densities with supports $\mathcal{X}_1, \ldots, \mathcal{X}_K$ and let $\boldsymbol{\alpha} = (\alpha_1, \ldots, \alpha_K)$ be a set of weights in the open $K$-simplex – i.e. $\alpha_i \geq 0$ and $\sum_{j=1}^{K} \alpha_j = 1$. Then, the logarithmically pooled density is

$$\pi_{\boldsymbol{\alpha}}(x) = t(\boldsymbol{\alpha}) \prod_{j=1}^{K} [f_j(x)]^{\alpha_j}, \tag{5}$$

where $t(\boldsymbol{\alpha})$ is a normalizing constant. This density combination ensures $\pi_{\boldsymbol{\alpha}}(\cdot)$ is unimodal on its support $(\cap_{j=1}^{K} \mathcal{X}_j)$. In log-space,

$$\log \pi_{\boldsymbol{\alpha}}(x) = \log\left(t(\boldsymbol{\alpha})\right) + \sum_{j=1}^{K} \alpha_j \log(f_j(x)). \tag{6}$$

The pool of log-normal distributions with parameters $\mu_i$ and $\sigma_i^2$ is a log-normal distribution with parameters defined according to equations (7) and (8) (Carvalho et al., 2023):

$$\mu^* = \frac{\sum_{i=0}^{K} w_i \mu_i}{\sum_{i=0}^{K} w_i}, \tag{7}$$

$$\sigma^{*2} = \left[\sum_{i=0}^{K} w_i\right]^{-1}, \tag{8}$$

where $w_i = \alpha_i / \sigma_i^{*2}$.

In contrast, a <u>linear</u> mixture would be

$$\bar{\pi}_{\boldsymbol{\alpha}}(x) = \sum_{j=1}^{K} \alpha_j f_j(x). \tag{9}$$

In log-space,

$$\log \bar{\pi}_{\boldsymbol{\alpha}}(x) = \log\left(\sum_{j=1}^{K} \alpha_j f_j(x)\right),$$

which can be evaluated using standard *log-sum-exp* techniques.

#### 2.2 Computing the ensemble models

Figure S6 shows the steps for constructing the ensemble models. The first step was to get the predictions submitted by the teams to the Mosqlimate platform, using *mosqlient*, a Python package designed to filter the data from the Mosqlimate API. Second, for each model prediction, we found the log-normal parameters $\mu$ and $\sigma$ – respectively mean and standard deviation in logspace – which lead to the best fit to the prediction intervals and median – step 2 in Figure S6.

The prediction of each model is then characterized by the percentiles $l$ (5th percentile), $m$ (50th percentile), and $u$ (95th percentile). Then we employ a numerical optimization method to determine the mean ($\mu^*$) and variance ($v^*$) of the log-normal distribution.

For cases where $m > 0$, the optimization problem is formulated as:

$$(\mu^\star, \sigma^\star) = \text{argmin}_{\mu \in \mathbb{R}, \sigma \in \mathbb{R}_+} \frac{|u - \hat{u}(\mu, \sigma)|}{u} + \frac{|m - \hat{m}(\mu, \sigma)|}{m}, \tag{10}$$

where $\hat{m}(\mu, \sigma)$ and $\hat{u}(\mu, \sigma)$ are the median and (upper) 95% quantile of a log-normal with parameters $\mu$ and $\sigma$.

For the special case where $m = 0$, the optimization problem becomes:

$$(\mu^\star, \sigma^\star) = \text{argmin}_{\mu \in \mathbb{R}, \sigma \in \mathbb{R}_+} \frac{|u - \hat{u}(\mu, \sigma)|}{u} \tag{11}$$

The routine was implemented in Python, using the *scipy* package and the *Nelder-Mead* optimization method and was initialized with $\mu^* = \log(m)$ and $v^* = 0.5$. The bounds for the algorithm were defined as $[-5 \cdot |\mu^*|, 5 \cdot |\mu^*|]$ for $\mu^*$, based on the initialized value of $\mu^*$, and $[0, 10]$ for the $v^*$ parameter.

After the fitting process, two ensemble models were constructed, where the first ensemble ($E_1$) is an equal-weight linear combination of the model predictions, whereas the second ensemble ($E_2$)uses logarithmic pooling to combine model predictions using weights optimized by minimizing the Continuous Ranked Probability Score (CRPS). We then computed the weights using the 2023 observed data, generating in-sample predictions for 2023, and used the same weights to generate out-of-sample predictions for the 2024 season.

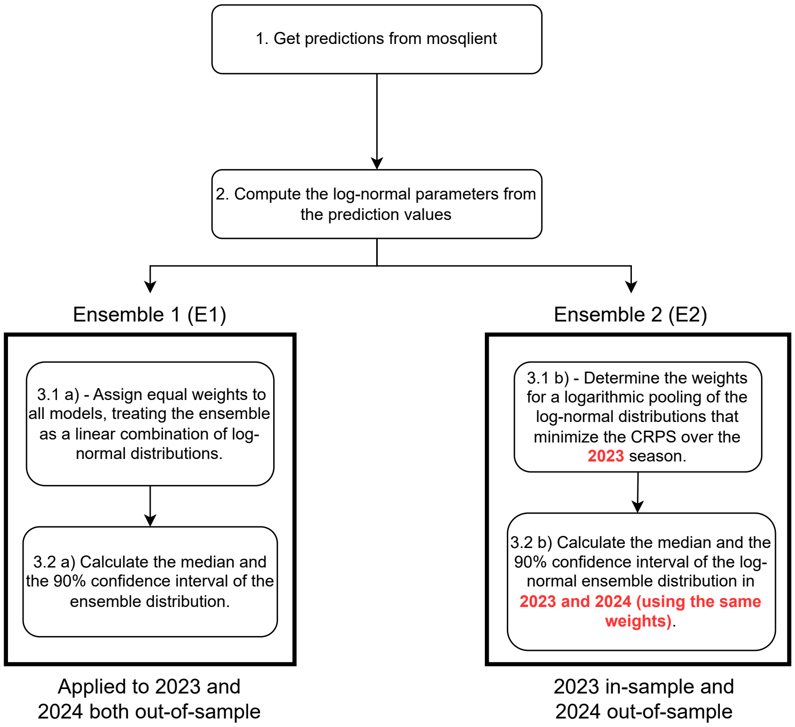


**Fig. S6.** Workflow to compute the ensemble models.

#### Ensemble $E_1$

The step 3.1 $(a)$ of Figure S6 was computed using equation 9, assuming $\alpha_i = \frac{1}{K}$ for each $i \in \{1, \ldots, K\}$. To compute the percentile values, the step 3.2 $(a)$ of Figure S6, the numerical method described below was used:

1. The density values were computed over the range from $10^{-6}$ to $10^5$, with $10^5$ points, and stored in a vector;
2. The CDF (cumulative distribution function) values were computed using the density values from the previous step and the composite trapezoidal rule utilizing the *scipy* package;
3. Interpolation was performed to compute the inverse CDF, allowing the calculation of the percentile values.

#### Ensemble $E_2$

The step 3.1 $b)$ of Figure S6 was computed using equation 7, and 8 to combine the log-normal distributions. To define the best weights, the equation 12 was minimized:

$$\text{argmin}_{\boldsymbol{\alpha}} \sum_{t=1}^{W} CRPS(\mu^*, v^*), \tag{12}$$

where $W$ is the number of weeks in the season, and CRPS is computed as in (13), implemented in *scoringrules*[1]:

$$\text{CRPS}(\log\mathcal{N}(\mu,\sigma), y) = y[2\Phi(y) - 1] - 2\exp\left(\mu + \frac{\sigma^2}{2}\right)\left[\Phi(\omega - \sigma) + \Phi(\frac{\sigma}{\sqrt{2}})\right], \tag{13}$$

where $\Phi$ is the CDF of the standard normal distribution and $\omega = \frac{\log y - \mu}{\sigma}$.

To compute the percentile values, the step 3.2 *b*) of Figure S6 it was used the percentile function of a log-normal distribution implemented in *scipy*(Virtanen et al., 2020).

### 3 Results: Score Values for Models and Ensembles

All models and ensembles were assessed using the three score metrics proposed: CRPS, interval score and log score.

The scores were computed per week, and subsequently averaged to produce a global score per season. In the following figures, for each pair (model, state) we show the average Interval (Figure S7) and Log scores (S8). The average CRPS table is found in the main text.

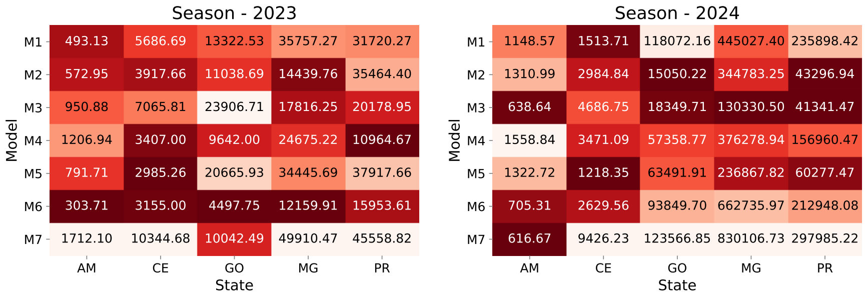


**Fig. S7.** Average interval score of models $M_1$ to $M_7$ predictions for season 2023 and 2024, at each mandatory state (lower values are better). Notice that the best model varies by year and state.

[1] https://github.com/frazane/scoringrules

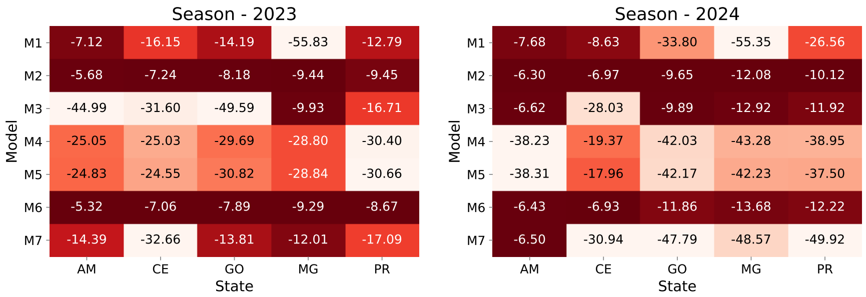


**Fig. S8.** Average log score of models $M_1$ to $M_7$ predictions for season 2023 and 2024, at each mandatory state (Larger values are better). Notice that the best model varies by year and state.

#### 3.1 Scores of the Ensemble models

Similar to the individual models, the scores were computed for the two Ensembles and are shown in figures S9, S10 and S11.

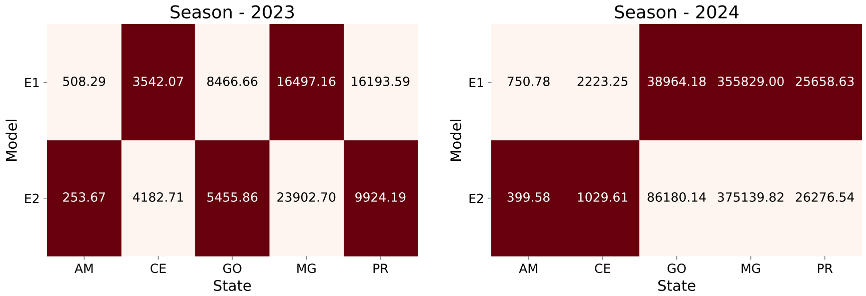


**Fig. S9.** Interval score values of the ensembles in season 2023 and 2024 for each mandatory state (Lower is better). The interval score values here are the average score for the entire prediction window.

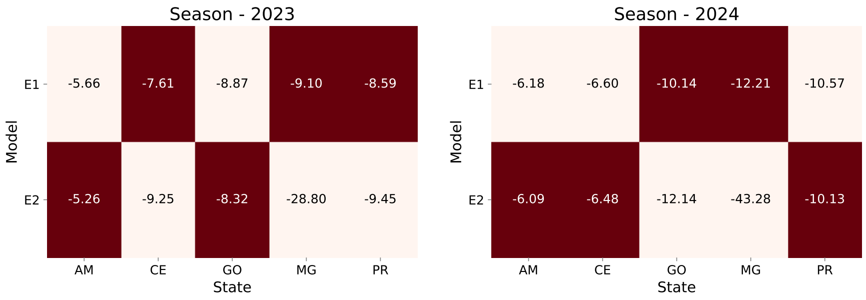


**Fig. S10.** Log score values of the ensemble in season 2023 and 2024 for each mandatory state (Larger is better). The log score values here are the average score for the entire prediction window.

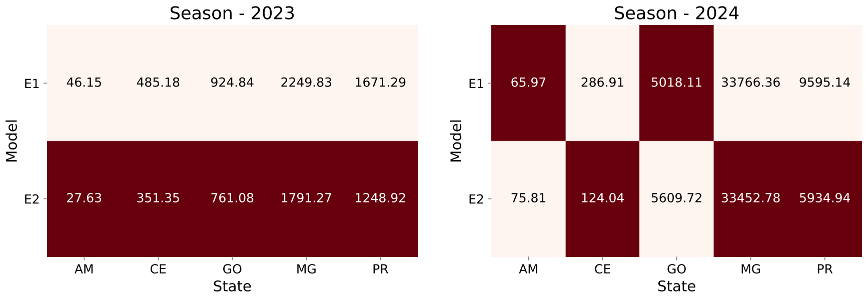


**Fig. S11.** CRPS score values of the ensembles in the season 2023 and 2024 for each mandatory state (Lower is better). The CRPS values here are the average score for the entire prediction window.

#### 3.2 Weekly Score Values of Individual and ensemble models

The weekly score values are relevant because they help us to understand the performance of the models over time. Figures S12 and S13 show the weekly CRPS scores for models and ensembles, respectively.

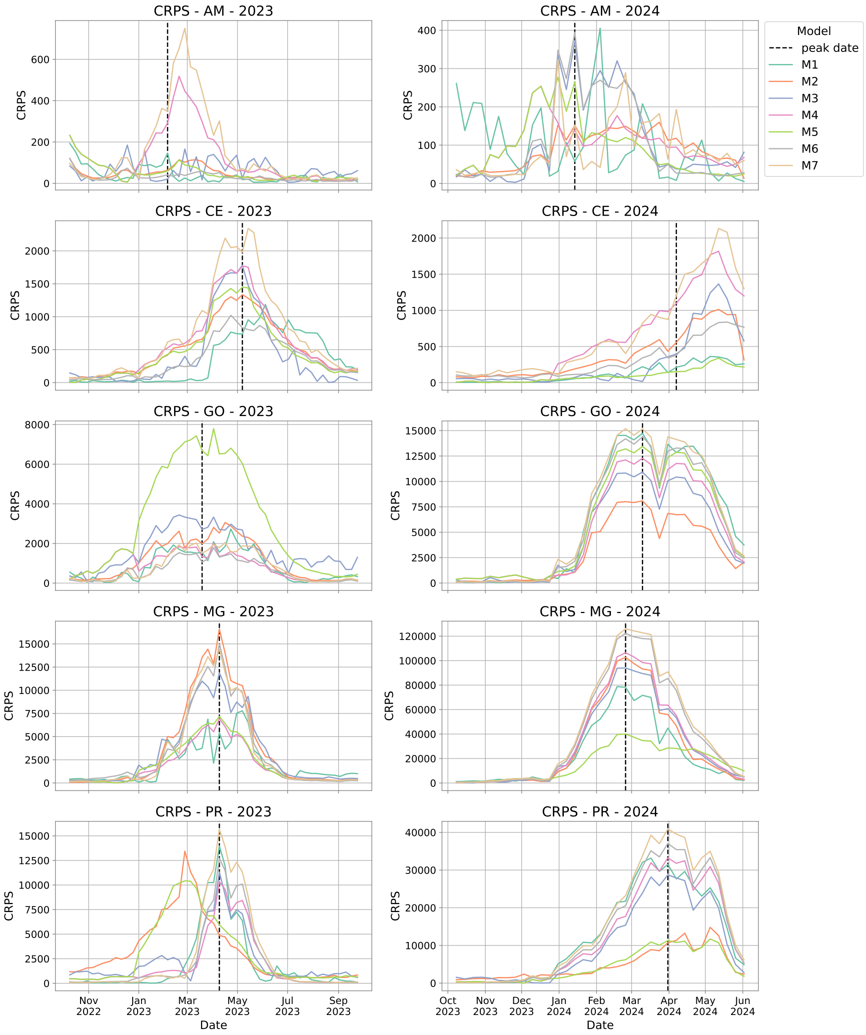


**Fig. S12.** CRPS values of all models in season 2023 and 2024 for each mandatory state (Lower is better). Notice that the best model varies by year and state. The CRPS values here are the weekly values.

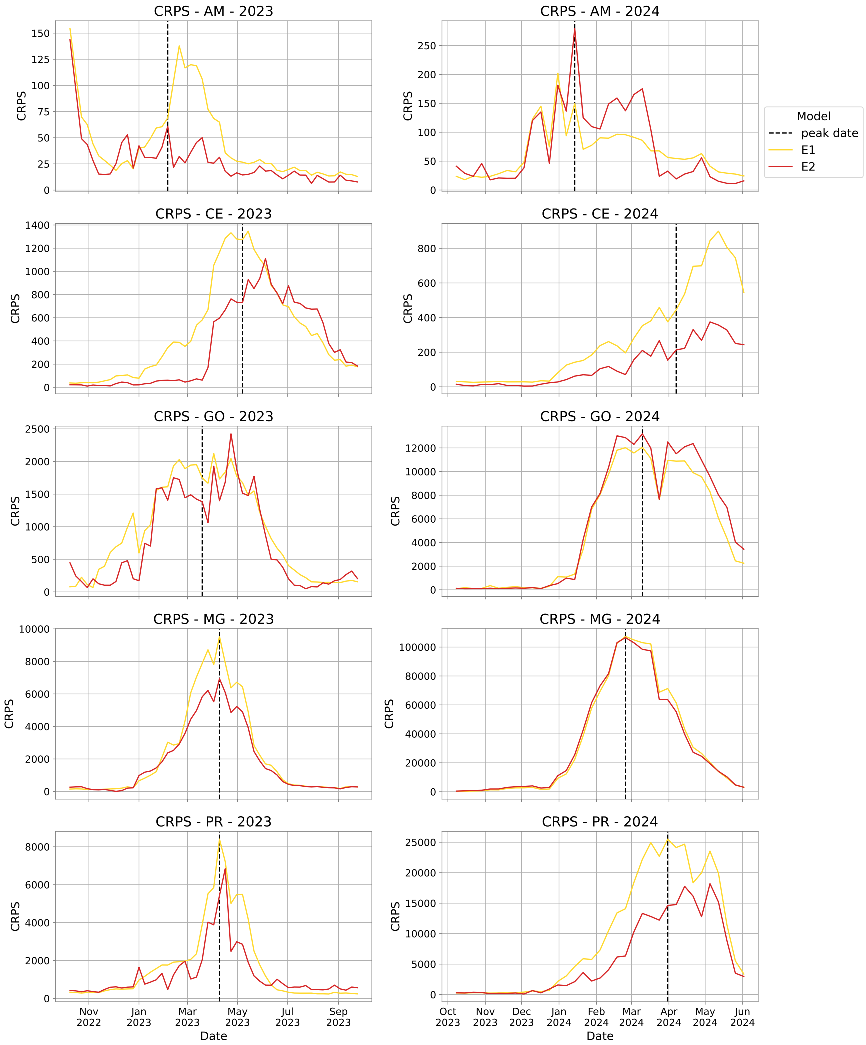


**Fig. S13.** CRPS values of ensembles in season 2023 and 2024 for each mandatory state (Lower is better). The CRPS values here are the weekly values.

Figures S14 and S15, show the weekly log scores for models and ensembles, respectively.

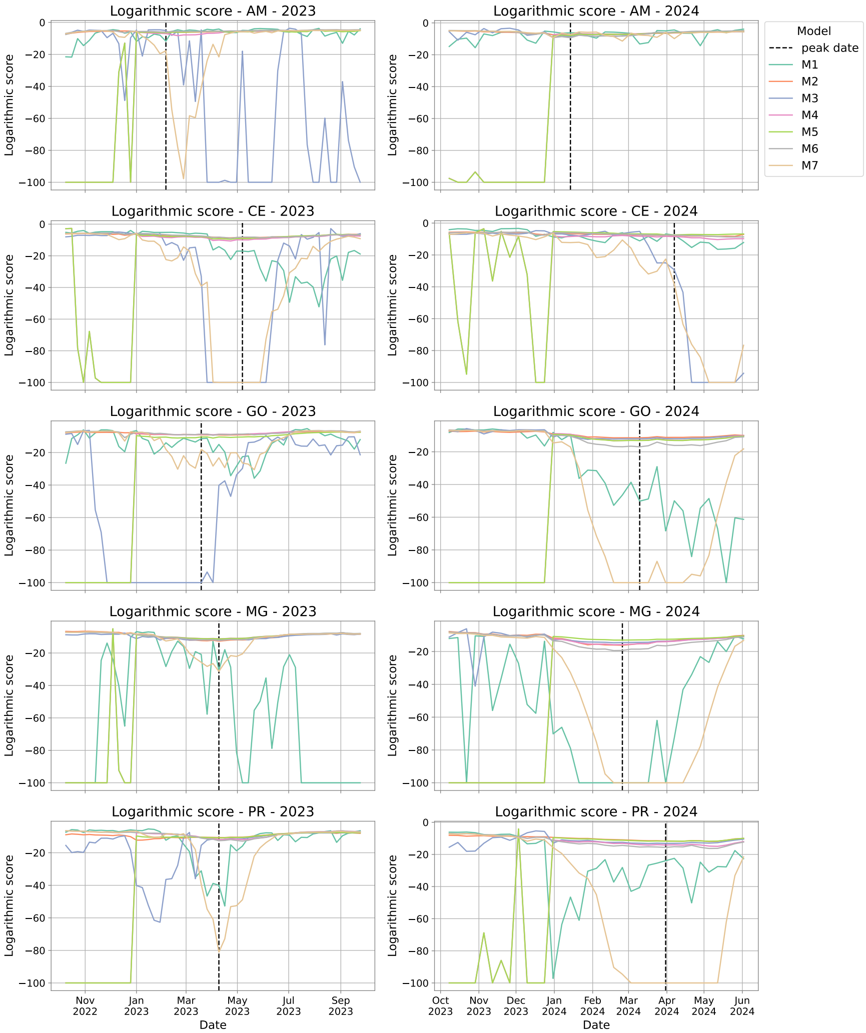


**Fig. S14.** Log score values of all models in season 2023 and 2024 for each mandatory state (Larger is better). Notice that the best model varies by year and state. The log score values here are the weekly values.

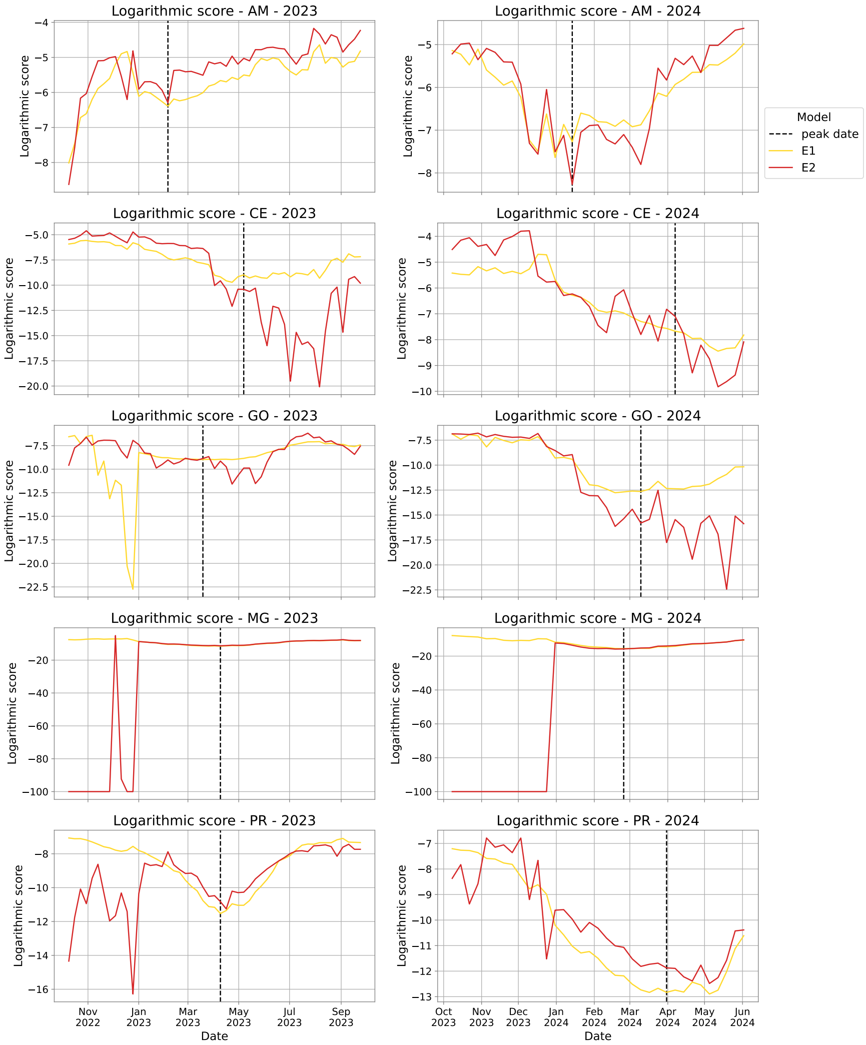


**Fig. S15.** Log score values of ensembles in season 2023 and 2024 for each mandatory state (Larger is better). Notice that the best model varies by year and state. The log score values here are the weekly values.

Figures S16 and S17, show the weekly interval scores for models and ensembles, respectively.

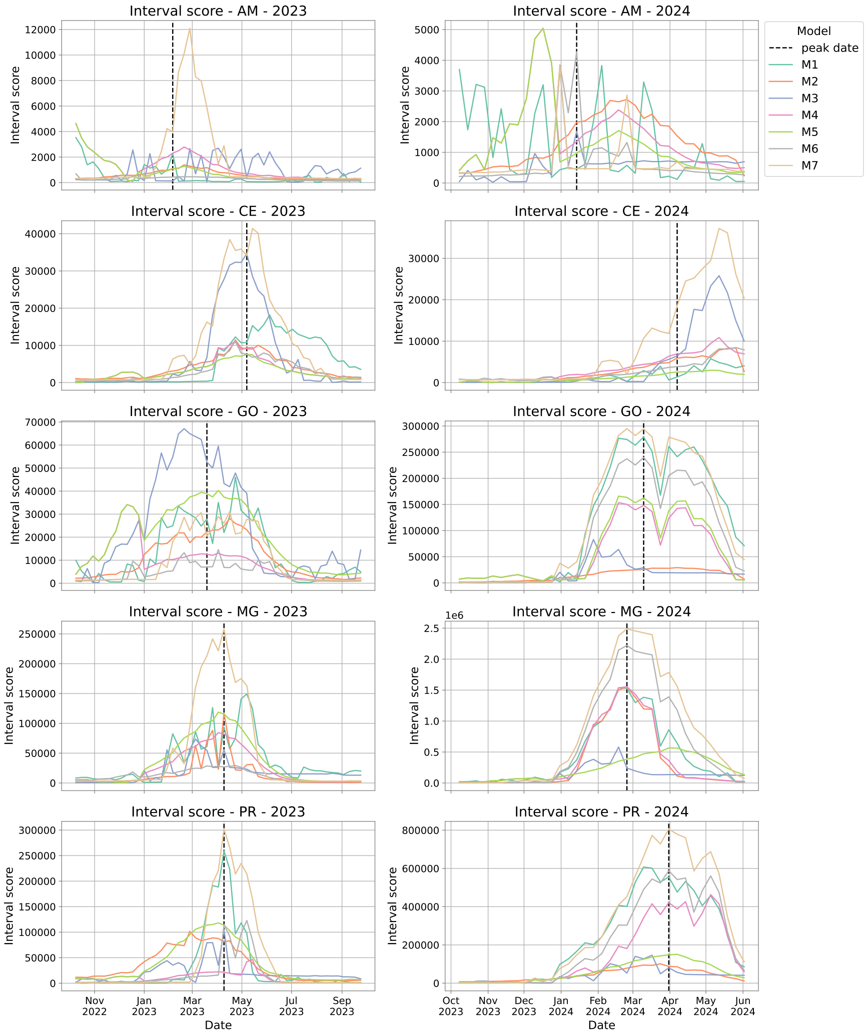


**Fig. S16.** Interval score values of all models in season 2023 and 2024 for each mandatory state (Lower is better). The interval score values here are the weekly values.

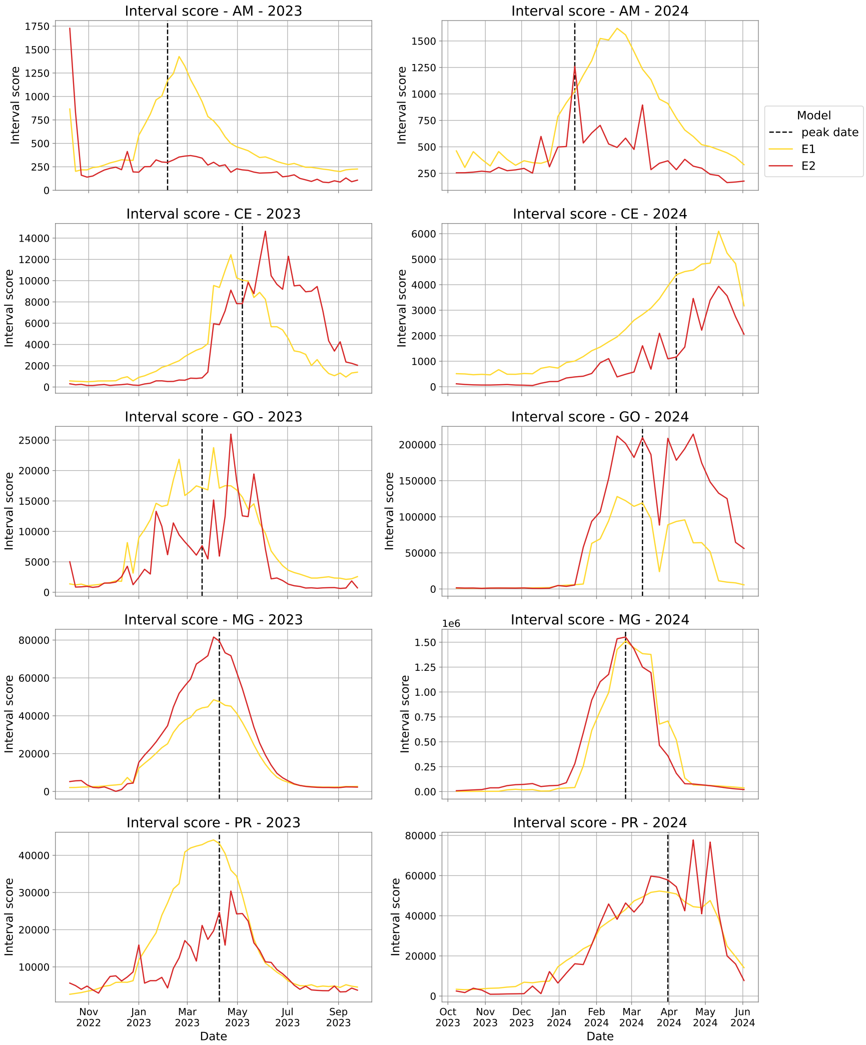


**Fig. S17.** Interval score values of ensembles in season 2023 and 2024 for each mandatory state (Lower is better). The interval score values here are the weekly values.

### 4 Sprint data and parameters description

This data dictionary has been developed for the Infodengue Sprint 2024: Predictive Modeling of Dengue in Brazil. It contains detailed documentation of the tables available for the Sprint, providing essential information for data analysis and modeling.

#### 4.1 Disease data

Period: epiweek 201001 to epiweek 2024241 Aggregation: cases aggregated by the epidemiological week of dengue symptom onset; municipality Sources: from SINAN and IBGE, organized by Infodengue

**Supplementary Table S2.** Description of 2024 dengue sprint case data.

| Name | Type | Description |
|---|---|---|
| date | YYYY-mm-dd | First day of the epiweek (Sunday) |
| year | int(YYYY) | Year |
| epiweek | int(YYYYWW) | Epidemiological week defined by the date of symptom onset |
| geocode | int | Municipality code (source IBGE) |
| casos | int | Number of cases per week, classified as probable dengue cases[2] |
| Rt | float | Point estimate of the reproductive number of dengue provided by Infodengue |
| p_rt1 | float | Probability (Rt ¿ 1) provided by Infodengue |
| regional* | str | Health district |
| regional_geocode | int | Health district code |
| macroregional | str | Health macroregion |
| macroregional_geocode | int | Health macroregion code |
| uf | str | Federative Unit (state) |
| train_1 | bool | Data for the first training (pre-season 22/23) |
| train_2 | bool | Data for the first validation (season 22/23) |
| target_1 | bool | Data for the second training (pre-season 23/23) |
| target_2 | bool | Data for the second validation (season 23/24) |

** Regional and Macroregional are the subdivisions used by the Brazilian Ministry of Health.*

#### 4.2 Climate data

Reanalysis hourly data from ERA5 provide spatial resolution of grids obtained from the Copernicus Climate Data Store. These data was summarized by week and city by the Mosqlimate project, covering period from 201001 to 202423. Period: epiweek 201001 to epiweek 2024234 Aggregation: temperature, humidity and precipitation, originally by hour, was first aggregated by day (min, max, mean), and these daily measures were aggregated by epidemiological week (mean). Sources: Copernicus Climate Data Store ERA5, organized by Mosqlimate.

**Supplementary Table S3.** Description of 2024 dengue sprint climate data.

| **Parameter Name** | **Type** | **Description** |
|---|---|---|
| date | YYYY-mm-dd | First day of the epiweek (Sunday) |
| epiweek | int (YYYYWW) | Epidemiological week |
| geocode | int | Municipality code |
| temp_min | float (°C) | Minimum temperature |
| temp_med | float (°C) | Mean temperature |
| temp_max | float (°C) | Maximum temperature |
| precip_min | float (mm/h) | Minimum precipitation rate |
| precip_med | float (mm/h) | Average precipitation rate |
| precip_max | float (mm/h) | Maximum precipitation rate |
| precip_tot | float (mm) | Total precipitation |
| pressure_min | float (atm) | Minimum daily sea level atmospheric pressure[3] |
| pressure_med | float (atm) | Average atmospheric pressure |
| pressure_max | float (atm) | Maximum atmospheric pressure |
| rel_humid_min | float (%) | Minimum relative humidity |
| rel_humid_med | float (%) | Average relative humidity |
| rel_humid_max | float (%) | Maximum relative humidity |
| thermal_range | float (°C) | Difference between daily maximum and minimum temperature averaged by week |
| rainy_days | int | Number of days in the week which ‘precip_tot ¿ 0.03‘. |

#### 4.3 Sea surface temperature and level oscillations data

These data were organized into three files contained:

**Supplementary Table S4.** Description of 2024 dengue sprint sea surface temperature and level oscillations data

| Parameter | Description |
|---|---|
| El Niño-Southern Oscillation | It is a climate pattern in the Pacific Ocean that has two phases: El Niño and La Niña. During an El Niño event, the trade winds weaken and warm, nutrient-poor waters are no longer pushed by the winds, and sea level rises in the eastern tropical Pacific and falls in the western tropical Pacific. La Niña is the opposite phase, with warm water piling up in the western Pacific and colder water in the eastern Pacific. This causes higher sea level in the western tropical Pacific and lower sea level in the eastern tropical Pacific. |
| The Indian Ocean Dipole - IOD | It is a climate pattern affecting the Indian Ocean. During a positive phase, warm waters are pushed to the Western part of the Indian Ocean, while cold deep waters are brought up to the surface in the Eastern Indian Ocean. This pattern is reversed during the negative phase of the IOD. |
| The Pacific Decadal Oscillation - PDO | It is a long-term (10-20 year) oscillation of the Pacific Ocean in response to atmospheric changes. During a warm (positive) phase, low atmospheric pressure over the Aleutian Islands causes ocean currents to bring warm waters to the Eastern Pacific Ocean and along the North American coast. Cool, nutrient-rich waters are displaced to the western Pacific Ocean. This leads to higher sea levels along the coast. |

#### 4.4 Environmental data

Environmental characteristics of the municipalities. Period: 2010 Sources: IBGE, Embrapa

**Supplementary Table S5.** Description of 2024 dengue sprint environmental characteristics of the municipalities data.

| Parameter | Type | Description |
|---|---|---|
| geocode | int | IBGE municipality code |
| muni_name | str | Municipality name |
| altitude | int | Altitude |
| Koppen | str | Main climate type |
| biome | str | Main biome type |

##### 4.5 Demographic data

Population data. Files with population by city and year (2010 - 2021) and municipalities that are regions of influence (REGIC).

**Supplementary Table S6.** Description of 2024 dengue sprint population, municipalities, and regions of influence data.

| Parameter | Type | Description |
|---|---|---|
| MUNIC_RES | int | Same as geocode in other tables |
| ANO | int | Year (YYYY) |
| POPULACAO | int | Population of the city |
| geocode | int | IBGE's municipality code |
| UF | str | Federative units (Brazil) |
| name_muni | str | Municipality name |
| hierarquia | str | Municipality Influence |
